# Determinants, Barriers, and Completion Patterns of Routine Childhood Immunization in Bayelsa State, Nigeria

**DOI:** 10.64898/2026.05.20.26353707

**Authors:** Vivian Ibienebakabobo Promise, Morufu Olalekan Raimi

**Affiliations:** Department of Public Health, Faculty of Health Sciences, Bayelsa Medical University, Yenagoa, Bayelsa State, Nigeria; Niger-Delta Institute for Emerging and Re-Emerging Infectious Diseases (NDIERID), Federal University Otuoke, Bayelsa State. Nigeria

**Keywords:** Vaccination adherence, Immunization completion, Dropout, Local Government Areas, Phone reminders, Childhood immunization, Nigeria, Public health

## Abstract

**Background:** Incomplete childhood vaccination undermines individual and herd immunity and increases vulnerability to vaccine-preventable diseases. Understanding local determinants of vaccination adherence is essential for targeted interventions. This study assessed routine immunization completion and dropout patterns among children aged 0-15 months in Bayelsa State, Nigeria.

**Objectives:** To determine vaccination completion rates, identify factors influencing adherence, analyze temporal patterns across immunization milestones, and provide evidence-based recommendations for improving coverage.

**Methods:** A comparative longitudinal study was conducted from March 2023 to July 2024 across three Local Government Areas (LGAs), representing each senatorial district. A total of 369 mother-child pairs (123 per LGA) were enrolled. Data were obtained from health facility immunization registers and supplemented with semi-structured questionnaires. Children were followed through the 6th week, 10th week, 14th week, 9th month, and 15th month immunization visits. Completion rates were analyzed using descriptive statistics and chi-square tests. Ethical approval was obtained from the State Ministry of Health, and informed consent was obtained from all mothers.

**Results:** Completion rates varied across LGAs, with the highest in LGA C (86.2%) and lowest in LGA B (61.0%). Phone-based reminders achieved the highest adherence, outperforming routine and home visit strategies. Progressive attrition was observed along the immunization schedule, with dropout exceeding completion by the 15th month. Principal reasons for non-completion included forgetfulness, travel, and caregiver busyness. Maternal age, education, and occupation significantly influenced adherence, indicating disparities across LGAs.

**Conclusion:** Vaccination adherence is shaped by maternal characteristics and operational strategies. While early-stage coverage is high, attrition increases at later milestones, particularly in LGAs with lower resource engagement.

**Recommendations:** Implement targeted phone-based reminders, milestone-specific outreach, and community engagement programs to reduce dropout, enhance timely completion, and strengthen childhood immunity.

## 1. Introduction

Immunization is a highly cost-effective public health intervention, preventing an estimated three million deaths annually and reducing the global burden of vaccine-preventable diseases through direct protection and herd immunity (World Health Organization, 2020; World Health Organization, 2021; United Nations Children’s Emergency Fund, 2023; Morufu *et al*., 2021; Raimi *et al*., 2021b). Despite these gains, achieving and sustaining ≥90% coverage, a threshold for optimal population-level protection, remains challenging in many low- and middle-income countries, where systemic inequities, weak health systems, and sociobehavioural barriers impede progress (World Health Organization, 2024; World Health Organization, 2025; Sarker *et al*., 2019; Tsegaw *et al*., 2024; Raimi *et al*., 2022). Immunization performance is increasingly recognized as a sensitive indicator of broader health system functionality, reflecting the interplay of service delivery capacity, community trust, and governance structures (World Health Organization Africa, 2019a; Kakwi *et al*., 2025; Oginifolunnia *et al*., 2025; Akabueze & Raimi, 2026; Ezekiel & Raimi, 2026). Vaccine hesitancy, defined as the delay or refusal of vaccination despite availability, further complicates uptake, shaped by misinformation, risk perception, and sociocultural factors (World Health Organization, 2014; Raimi *et al*., 2021a; Kakwi *et al*., 2024a, b; Babbo *et al*., 2025; Elemuwa *et al*., 2024a). Consequently, understanding determinants of completion, not just initiation, is critical to advancing effective immunization strategies (Raimi *et al*., 2019). High rates of zero-dose and partially immunized children, especially among those aged 12-23 months, undermine cumulative schedule efficacy and erode herd immunity, sustaining vulnerability to outbreaks and avoidable mortality (Tsegaw *et al*., 2024; Sarker *et al*., 2019; World Health Organization, 2024; Promise *et al*., 2024; Raimi, 2025a-d). Globally, 14.5 million children remain zero-dose and 6.5 million partially immunized, disproportionately concentrated in countries such as Nigeria (World Health Organization, 2024; United Nation Children Emergency Fund, 2023; World Health Organization, 2023; Mordecai *et al*., 2024; Oweibia *et al*., 2024). Drivers of non-completion differ from those of non-initiation and include logistical constraints, caregiver fatigue, opportunity costs, and health system inefficiencies (Sarker *et al*., 2019; Tsegaw *et al*., 2024; Abdulraheem *et al*., 2025a-c; Babbo *et al*., 2025). Incomplete immunization is often obscured in routine coverage metrics, masking critical dropout points along the vaccination continuum (Promise *et al*., 2026; Raimi *et al*., 2022; Kakwi *et al*., 2025; Oginifolunnia *et al*., 2025; Raimi, 2025c). Understanding these patterns is essential to disentangle layered determinants and guide interventions.

In Nigeria, these challenges are particularly pronounced. Despite sustained policy attention and intervention programs, the country continues to account for a significant proportion of under-immunized children, with regional disparities and high dropout rates between initial and subsequent doses (United Nation Children Emergency Fund, 2023; World Health Organization, 2024; Promise *et al*., 2024; Okechukwu *et al*., 2024; Uchenna *et al*., 2024). National surveys indicate that approximately one-third of children receiving the first pentavalent dose fail to complete the third, reflecting discontinuities in service utilization (Promise *et al*., 2024; World Health Organization, 2024; Raimi *et al*., 2022; Kakwi *et al*., 2024a; Raimi, 2025b; Adiama *et al*., 2025; Adaka *et al*., 2026). Bayelsa State exemplifies these challenges, with historically low routine immunization performance, dropout rates exceeding 30%, and persistent structural and behavioural barriers (World Health Organization Africa, 2019a; Promise *et al*., 2024; Okechukwu *et al*., 2024; Raimi *et al*., 2022; Aziba-anyam & Raimi, 2025; Promise *et al*., 2026; United Nation Children Emergency Fund, 2023; World Health Organization, 2025; Ogar *et al*., 2026; Elemuwa *et al*., 2024b). While donor-supported interventions and community-based strategies have produced incremental improvements, dropout remains substantial, raising questions about the sustainability and contextual adaptability of existing programs. Theoretically, health behaviour frameworks, such as the Health Belief Model, posit that vaccination adherence is influenced by perceived susceptibility, perceived benefits, and modifiable cues to action; however, empirical applications in high-burden Nigerian settings remain limited (World Health Organization, 2014; Raimi *et al*., 2021a; Kakwi *et al*., 2025; Abdulraheem *et al*., 2025a-c; Babbo *et al*., 2025). Emerging interventions, including mobile phone reminders and home visits, show promise but require further contextual validation (Promise *et al*., 2026; Oyo-Ita *et al*., 2021; Akabueze & Raimi, 2026; Ezekiel & Raimi, 2026; Raimi *et al*., 2022). This study addresses the evidence gap between initiation and completion of childhood immunization in Bayelsa State. Specifically, it aims to quantify incomplete immunization, identify its key determinants, and assess the influence of socio-demographic, behavioural, and health system factors on completion rates among children under two years.

## 2. Methods

### 2.1 Study setting

This study was conducted in Bayelsa State, Nigeria, located in the Niger Delta region of South-South Nigeria, a setting characterized by complex riverine and estuarine geographies that shape access to primary healthcare services and immunization delivery (Kemi, 2017; NGEX, 2013; World Health Organization Africa, 2019a; Raimi *et al*., 2022; Tinimoye *et al*., 2026). The state comprises eight Local Government Areas (LGAs), each subdivided into political wards containing communities with functional primary healthcare centres responsible for routine immunization programmes targeting children under two years of age (National Population Commission, 2006; Raimi *et al*., 2019; Oyo-Ita *et al*., 2021; Elemuwa *et al*., 2024b; Oginifolunnia *et al*., 2025). Although Bayelsa is among the least populous states in Nigeria, with an estimated projected population exceeding two million in 2023, its dispersed settlement patterns and difficult terrain present unique challenges for equitable vaccine delivery and uptake (National Population Commission, 2006; United Nation Children Emergency Fund, 2023; World Health Organization, 2024; Raimi *et al*., 2022; Adelekun *et al*., 2025). Consistent with national planning assumptions, approximately 7.2% of the population constitutes children under two years of age, and this proportion is routinely disaggregated and allocated across health facilities as target populations for immunization monitoring and evaluation (National Population Commission, 2006; World Health Organization, 2021; Promise *et al*., 2024; Olulaja & Jaber, 2026; Raimi *et al*., 2019). This structured distribution underpins service delivery planning but may obscure micro-level disparities in access and utilization, particularly in hard-to-reach riverine communities.

### 2.2 Study design

A comparative longitudinal study design was employed to enable systematic follow-up of immunization trajectories among children under two years of age across selected health facilities. This design permits the temporal assessment of vaccine uptake and completion patterns, thereby capturing dropout dynamics that are often overlooked in cross-sectional surveys (Tsegaw *et al*., 2024; Sarker *et al*., 2019; Landoh *et al*., 2016; Raimi *et al*., 2022; Promise *et al*., 2026). Longitudinal approaches are particularly advantageous in immunization research, where adherence to multi-dose schedules is contingent upon sustained caregiver engagement and consistent health system performance (Lakew *et al*., 2015; Mekonnen *et al*., 2021; Domek *et al*., 2019; Chen *et al*., 2016; Eze *et al*., 2018). Furthermore, this design aligns with emerging evidence advocating for real-time monitoring and adaptive intervention strategies to improve vaccine completion rates in resource-constrained settings (Raimi *et al*., 2022; Akabueze & Raimi, 2026; Ezekiel & Raimi, 2026; Kakwi *et al*., 2025; Oginifolunnia *et al*., 2025). By leveraging routine health facility records alongside primary data collection, the study integrates both retrospective and prospective elements, thereby enhancing the robustness and validity of the findings.

### 2.3 Study population and eligibility criteria

The study population comprised children aged 0-15 months and their mothers or primary caregivers residing within the selected communities. Both immunization-naïve and partially immunized children were included to capture full coverage trajectories.

#### 2.3.1 Inclusion and Exclusion Criteria

##### Inclusion criteria

i. Children aged 0-15 months registered in the selected health facilities’ immunization registers.
ii. Mothers or caregivers who provided informed consent.

##### Exclusion criteria

i. Children with medical contraindications to vaccination.
ii. Caregivers unwilling to participate or unavailable for follow-up visits.
iii. Children residing outside the selected communities for the study period (Promise *et al*., 2026; Okechukwu *et al*., 2024).

### 2.4 Sample size determination

The minimum sample size of 365 was calculated using Cochran’s formula for single proportions, incorporating a 10% non-response adjustment to account for potential attrition and incomplete data. The calculation assumed a prevalence (p) of 31% based on prior immunization coverage estimates, with a standard normal deviate (Z) of 1.96 corresponding to a 95% confidence level and a margin of error (d) of 0.05 (MIC/NIC, 2022; Landoh *et al*., 2016; Sarker *et al*., 2019; Tsegaw *et al*., 2024; Promise *et al*., 2024). This methodological approach is widely adopted in public health research to ensure adequate statistical power and precision in estimating population parameters (Abdur *et al*., 2019; Lakew *et al*., 2015; Raimi *et al*., 2019; Olulaja & Jaber, 2026; Babbo *et al*., 2025). The formula is expressed as:

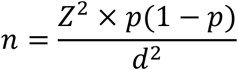

where *n* represents the required sample size, *Z* the standard normal deviate, *p* the estimated proportion, and *d* the desired level of precision. The inclusion of a non-response margin enhances the reliability of the study by mitigating potential biases arising from incomplete participation.

### 2.5 Sampling Procedure

Three LGAs were purposively selected, one from each senatorial district. Within each LGA, four wards were randomly selected, and one community per ward was chosen using simple random sampling. Within selected communities, health facilities providing routine immunization were identified, and all mother-child pairs meeting inclusion criteria were listed. Using systematic random sampling, 123 mother-child pairs per LGA were enrolled, yielding a total sample of 369 pairs. Selection bias was minimized by ensuring proportional representation from each ward and cross-verifying register data (Raimi *et al*., 2022; Kakwi *et al*., 2025).

### 2.6 Study Instrument

A semi-structured questionnaire was developed based on existing validated tools and the routine immunization schedule. The questionnaire was pilot-tested on 20 mother-child pairs in a non-study LGA to assess clarity, reliability, and comprehension. Revisions were made to improve wording, sequence, and cultural relevance. Data from facility registers were reconciled with caregiver interviews, and discrepancies were resolved through verification calls or repeated visits (World Health Organization, 2024; Promise *et al*., 2026).

### 2.7 Data Collection Procedure

Data were collected from both immunization registers and caregiver interviews at baseline and follow-up visits. Trained data collectors cross-checked register entries against caregiver recall to ensure accuracy. Missing data were documented, and imputation was performed for variables with ≤5% missingness; variables with higher missingness were analyzed separately and discussed as limitations (Sarker *et al*., 2019; Abdulraheem *et al*., 2025a). Phone-based and home visit follow-ups were conducted to track adherence across all scheduled immunization milestones.

### 2.8 Sampling technique

A multistage sampling technique was employed to ensure representativeness across the diverse geographical and administrative units of the state. In the first stage, one Local Government Area was selected from each of the three senatorial districts using simple random sampling, thereby capturing regional variability in immunization performance (Oyo-Ita *et al*., 2021; Raimi *et al*., 2022; Elemuwa *et al*., 2024b; Tinimoye *et al*., 2026; Promise *et al*., 2024). In the second stage, four wards were randomly selected from each chosen LGA, yielding a total of twelve wards. Subsequently, one community was randomly selected from each ward in the third stage, ensuring coverage of both rural and semi-urban contexts (National Population Commission, 2006; Raimi *et al*., 2019; Adelekun *et al*., 2025; Kakwi *et al*., 2025; Oginifolunnia *et al*., 2025). This hierarchical sampling framework enhances external validity while minimizing selection bias, particularly in settings characterized by spatial heterogeneity in healthcare access (Lakew *et al*., 2015; Sarker *et al*., 2019; Tsegaw *et al*., 2024; Raimi *et al*., 2022; Olulaja & Jaber, 2026).

### 2.9 Data collection and instruments

Data were collected using an interviewer-administered approach to enhance accuracy and completeness, particularly in populations with varying literacy levels. A semi-structured questionnaire adapted from the WHO Strategic Advisory Group of Experts (SAGE) vaccine hesitancy tool was utilized to capture socio-demographic characteristics, caregiver perceptions, and behavioural determinants of immunization uptake (World Health Organization, 2014; Adelekun *et al*., 2025; Kakwi *et al*., 2024b; Babbo *et al*., 2025; Raimi *et al*., 2021a). Evidence of vaccine uptake and completion was extracted from health facility immunization registers, thereby providing objective validation of self-reported data (Oginifolunnia *et al*., 2025; Promise *et al*., 2024; Raimi *et al*., 2019; Uchenna *et al*., 2024; Okechukwu *et al*., 2024). The integration of primary and secondary data sources strengthens measurement validity and reduces recall bias, a common limitation in immunization research (Landoh *et al*., 2016; Sarker *et al*., 2019; Tsegaw *et al*., 2024; Mekonnen *et al*., 2021; Domek *et al*., 2019).

### 2.10 Data management and statistical analysis

Data were entered and cleaned using Microsoft Excel® (version 2019) and subsequently analysed using Statistical Product and Service Solutions (SPSS) version 25. Descriptive statistics were computed to summarize categorical variables as frequencies and percentages, with results presented in tables and graphical formats. Inferential analysis was conducted using the chi-square (χ^2^) test to examine associations between categorical variables and immunization outcomes. Statistical significance was set at a p-value ≤0.05 at a 95% confidence interval. This analytical approach is consistent with established epidemiological methods for assessing relationships between predictors and categorical health outcomes in population-based studies.

### 2.11 Ethical considerations

Ethical approval for the study was obtained from the Bayelsa State Health Research Ethics Committee (BSHREC) [Approval No. BSHREC/2023/012]. The study adhered to international ethical standards for research with human participants, including respect for persons, beneficence, and justice. Written informed consent was obtained from all participants or, where applicable, from their caregivers prior to data collection. Participants (and caregivers) were informed that participation was voluntary, that they could withdraw at any time without penalty, and that confidentiality and anonymity would be maintained. No personally identifiable information was collected or disclosed, and all data were used solely for research purposes. Beyond its scientific contribution, the study aims to inform policy and practice by identifying gaps in immunization completion and providing evidence to strengthen public health interventions at the community, state, and national levels.

## 3. Results

A total of 369 mother-child pairs were included in the analysis, with equal representation from LGA A, LGA B, and LGA C (n = 123 each) (Table 1). In LGA A, male children accounted for 57.72% (71/123), while females constituted 42.28% (52/123). In contrast, LGA B had a higher proportion of female children (57.72%, 71/123) compared to males (42.28%, 52/123). In LGA C, the distribution was relatively balanced, with males representing 48.78% (60/123) and females 51.22% (63/123). Maternal age distribution showed that the majority of mothers were aged 20-29 years across all LGAs: 49.59% (61/123) in LGA A, 50.41% (62/123) in LGA B, and 46.34% (57/123) in LGA C. Mothers aged 30–39 years accounted for 35.77% (44/123) in LGA A and 29.27% (36/123) in both LGA B and LGA C. Adolescents (≤19 years) comprised 7.32% (9/123) in LGA A, 8.13% (10/123) in LGA B, and 7.32% (9/123) in LGA C. Mothers aged 40-49 years represented 7.32% (9/123) in LGA A, 12.20% (15/123) in LGA B, and 17.07% (21/123) in LGA C. Regarding educational attainment, secondary education was the most common level across all LGAs, reported by 55.28% (68/123) in LGA A and 52.85% (65/123) in both LGA B and LGA C. Primary education was reported by 26.83% (33/123) in LGA A, 35.77% (44/123) in LGA B, and 30.89% (38/123) in LGA C. Tertiary education was highest in LGA A at 17.89% (22/123), compared with 5.69% (7/123) in LGA B and 10.57% (13/123) in LGA C. No formal education was reported in LGA B and LGA C at 5.69% (7/123) each, while none was recorded in LGA A. Most respondents were married: 69.11% (85/123) in LGA A, 58.54% (72/123) in LGA B, and 65.04% (80/123) in LGA C. Occupational distribution indicated that business/trading was the predominant occupation across all LGAs, accounting for 53.66% (66/123) in LGA A, 38.21% (47/123) in LGA B, and 46.34% (57/123) in LGA C. Farming was reported by 23.58% (29/123) in LGA A, 26.83% (33/123) in LGA B, and 19.51% (24/123) in LGA C. Other occupations included students (13.01%, 16/123; 8.13%, 10/123; and 9.76%, 12/123 in LGA A, B, and C respectively), housewives (4.07%, 5/123; 4.07%, 5/123; and 3.25%, 4/123), and smaller proportions in artisan, civil service, fishing, hairdressing, tailoring, and teaching categories. Child birth order showed that first-born children constituted 47.15% (58/123) in LGA A, 38.21% (47/123) in LGA B, and 34.96% (43/123) in LGA C. Second-born children accounted for 25.20% (31/123) in LGA A and LGA C, and 24.39% (30/123) in LGA B. Third-born children were 8.13% (10/123) in LGA A, 11.38% (14/123) in LGA B, and 13.01% (16/123) in LGA C, while children of fourth birth order or higher represented 19.51% (24/123), 26.02% (32/123), and 26.83% (33/123) in LGA A, B, and C, respectively.

**Table 1.** Demographic characteristics of study participants across three local government areas in Bayelsa State, Nigeria.

| **Characteristic** | **LGA A (n = 123), n (%)** | **LGA B (n = 123), n (%)** | **LGA C (n = 123), n (%)** |
| --- | --- | --- | --- |
| **Child's gender** |  |  |  |
| Male | 71 (57.72) | 52 (42.28) | 60 (48.78) |
| Female | 52 (42.28) | 71 (57.72) | 63 (51.22) |
| **Mother's age (years)** |  |  |  |
| ≤19 | 9 (7.32) | 10 (8.13) | 9 (7.32) |
| 20–29 | 61 (49.59) | 62 (50.41) | 57 (46.34) |
| 30–39 | 44 (35.77) | 36 (29.27) | 36 (29.27) |
| 40–49 | 9 (7.32) | 15 (12.20) | 21 (17.07) |
| **Mother's highest level of education** |  |  |  |
| No formal education | 0 (0.00) | 7 (5.69) | 7 (5.69) |
| Primary | 33 (26.83) | 44 (35.77) | 38 (30.89) |
| Secondary | 68 (55.28) | 65 (52.85) | 65 (52.85) |
| Tertiary | 22 (17.89) | 7 (5.69) | 13 (10.57) |
| **Marital status** |  |  |  |
| Married | 85 (69.11) | 72 (58.54) | 80 (65.04) |
| Not married | 38 (30.89) | 51 (41.46) | 43 (34.96) |
| **Mother's occupation** |  |  |  |
| Artisan | 2 (1.63) | 0 (0.00) | 0 (0.00) |
| Civil servant | 5 (4.07) | 0 (0.00) | 2 (1.63) |
| Farming | 29 (23.58) | 33 (26.83) | 24 (19.51) |
| Fishing | 0 (0.00) | 4 (3.25) | 12 (9.76) |
| Hairdressing | 0 (0.00) | 14 (11.38) | 7 (5.69) |
| Housewife | 5 (4.07) | 5 (4.07) | 4 (3.25) |
| Student | 16 (13.01) | 10 (8.13) | 12 (9.76) |
| Tailor | 0 (0.00) | 9 (7.32) | 5 (4.07) |
| Teaching | 0 (0.00) | 1 (0.81) | 0 (0.00) |
| Business/Trading | 66 (53.66) | 47 (38.21) | 57 (46.34) |
| **Child's position** |  |  |  |
| 1st | 58 (47.15) | 47 (38.21) | 43 (34.96) |
| 2nd | 31 (25.20) | 30 (24.39) | 31 (25.20) |
| 3rd | 10 (8.13) | 14 (11.38) | 16 (13.01) |

| Characteristic | LGA A (n = 123), n (%) | LGA B (n = 123), n (%) | LGA C (n = 123), n (%) |
| --- | --- | --- | --- |
| ≥4th | 24 (19.51) | 32 (26.02) | 33 (26.83) |

*Data are presented as number (percentage). Percentages are calculated within each LGA.*

Figure 1 presents vaccination completion data across three LGAs: Ogbia (LGA A), Southern Ijaw (LGA B), and Sagbama (LGA C), under three follow-up strategies: routine control, home visits, and phone reminders. Across all LGAs, phone-based reminders consistently yielded the highest completion rates (LGA A: 106 completed, 17 not completed; LGA B: 98 completed, 20 not completed; LGA C: 97 completed, 19 not completed), whereas routine control measures and home visits resulted in lower completion rates. Non-completion was most pronounced in the control groups, highlighting gaps in adherence under passive or standard care. These data indicate that proactive engagement strategies, particularly phone follow-ups, substantially improve vaccination adherence across diverse settings. The consistent pattern across all LGAs suggests that communication-based interventions are both effective and scalable, providing actionable evidence for optimizing immunization programs in resource-limited contexts. Counts of individuals completing or not completing vaccination are presented for clarity, and the multi-panel format allows direct comparison of strategies and localities while maintaining a clean, legible layout suitable for publication.

**Figure 1:**
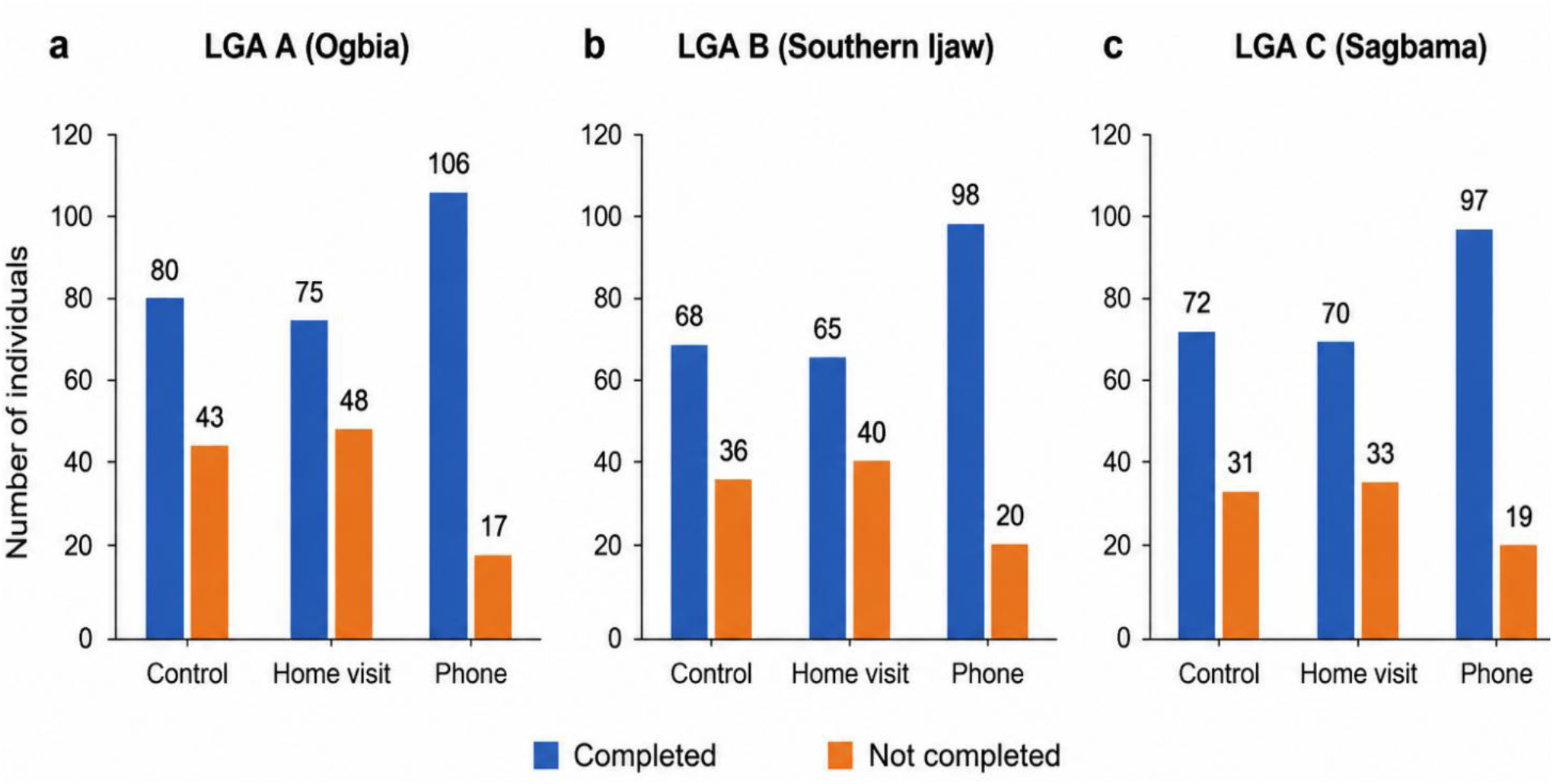
Vaccination coverage by follow-up strategy across Local Government Areas (LGAs)

Table 2 and Figure 2 show vaccination completion varied across the three Local Government Areas (LGAs). In LGA A, 65.0% (80/123) of children completed vaccination, while 35.0% (43/123) did not complete the recommended schedule. In LGA B, 61.0% (75/123) of children completed vaccination, compared with 39.0% (48/123) who did not. In contrast, LGA C recorded the highest completion rate, with 86.2% (106/123) of children completing vaccination and 13.8% (17/123) not completing. A chi-square test of independence indicated a statistically significant association between LGA and vaccination completion status (χ^2^ = 21.75, p = 0.001), suggesting that vaccination completion differed significantly across the three LGAs.

**Table 2.**
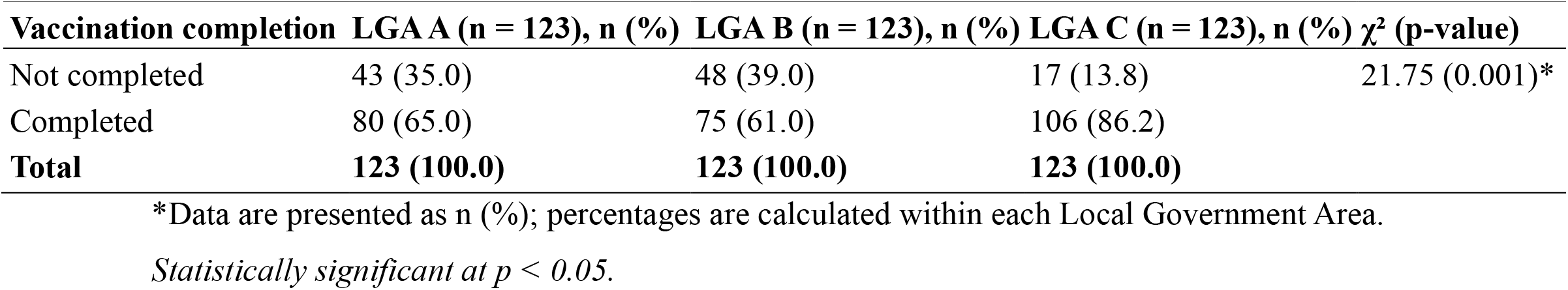
Comparison of vaccination completion across Local Government Areas.

| Vaccination completion | LGA A (n = 123), n (%) | LGA B (n = 123), n (%) | LGA C (n = 123), n (%) | $\chi^2$ (p-value) |
| --- | --- | --- | --- | --- |
| Not completed | 43 (35.0) | 48 (39.0) | 17 (13.8) | 21.75 (0.001)* |
| Completed | 80 (65.0) | 75 (61.0) | 106 (86.2) |  |
| **Total** | **123 (100.0)** | **123 (100.0)** | **123 (100.0)** |  |

\*Data are presented as n (%); percentages are calculated within each Local Government Area.
Statistically significant at $p < 0.05$ .

**Figure 2:**
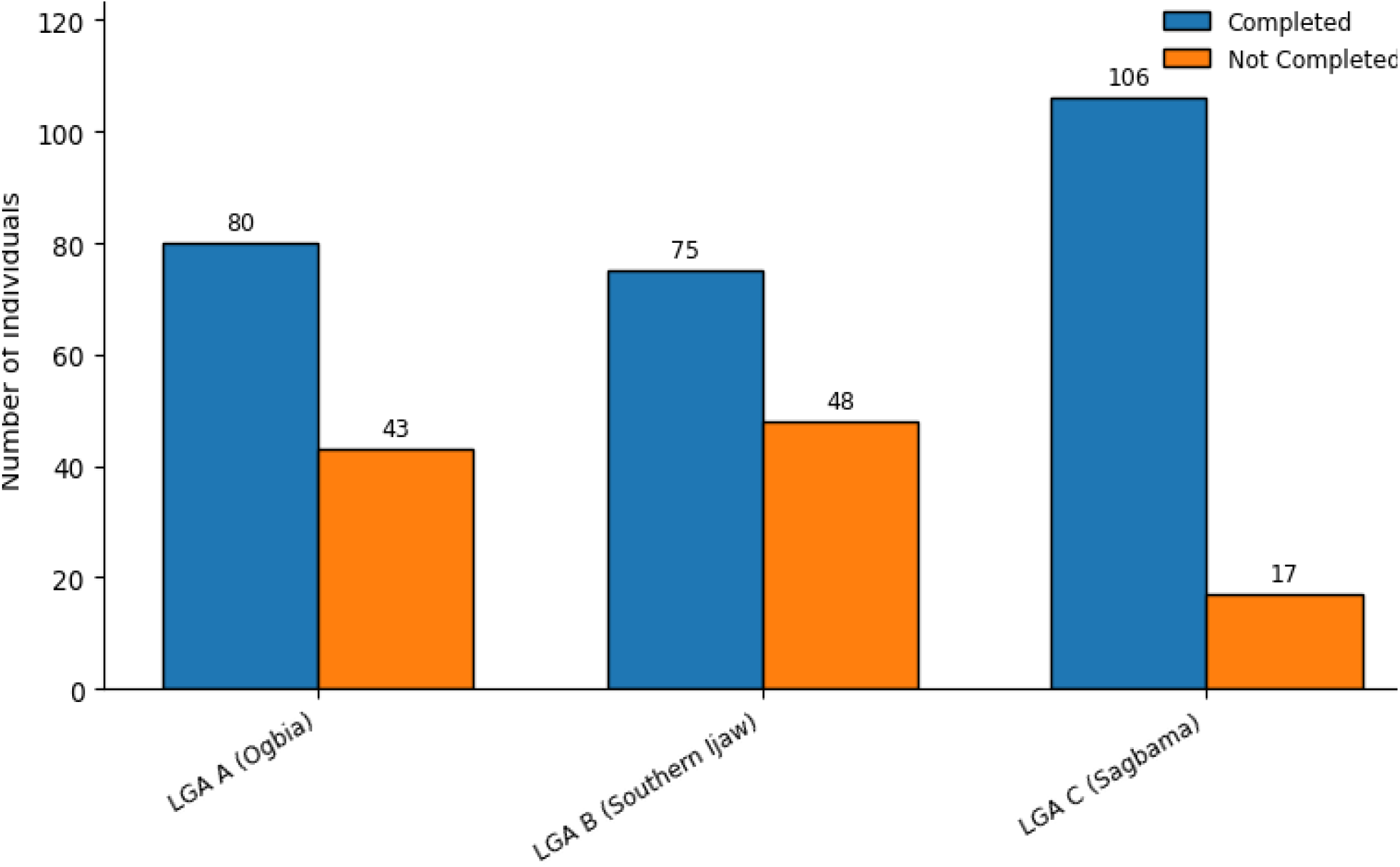
Vaccination completion across the three LGAs.

Figure 3 illustrates the distribution of vaccination completion and non-completion across key immunization milestones (6th week, 10th week, 14th week, 9th month, and 15th month) aggregated across the three study sites (LGAs). Overall, the frequency of completed vaccinations was highest at the early stages of the immunization schedule and progressively declined over time, while non-completion showed an inverse pattern. At the 6th week, completed vaccinations were highest (n = 293), with comparatively fewer cases of non-completion (n = 76). This trend persisted at the 10th week, where completion remained high (n = 284) relative to non-completion (n = 85). By the 14th week, a gradual decline in completion was observed (n = 262), accompanied by an increase in non-completion (n = 107). A more pronounced divergence emerged at the 9th month milestone, where completed vaccinations further declined to 226, while non-completion increased substantially to 143. This pattern culminated at the 15th month, where non-completion exceeded completion, with 291 cases of incomplete vaccination compared to only 78 completed cases. Figure 3 demonstrates a clear attrition pattern along the immunization continuum, characterized by high initial uptake followed by a progressive drop-off at subsequent visits, particularly at later milestones. This temporal distribution highlights critical points within the vaccination schedule where loss to follow-up is most pronounced. Figure 3 represents frequencies (n) of children who completed or did not complete vaccination at each scheduled immunization milestone across the three LGAs.

**Figure 3:**
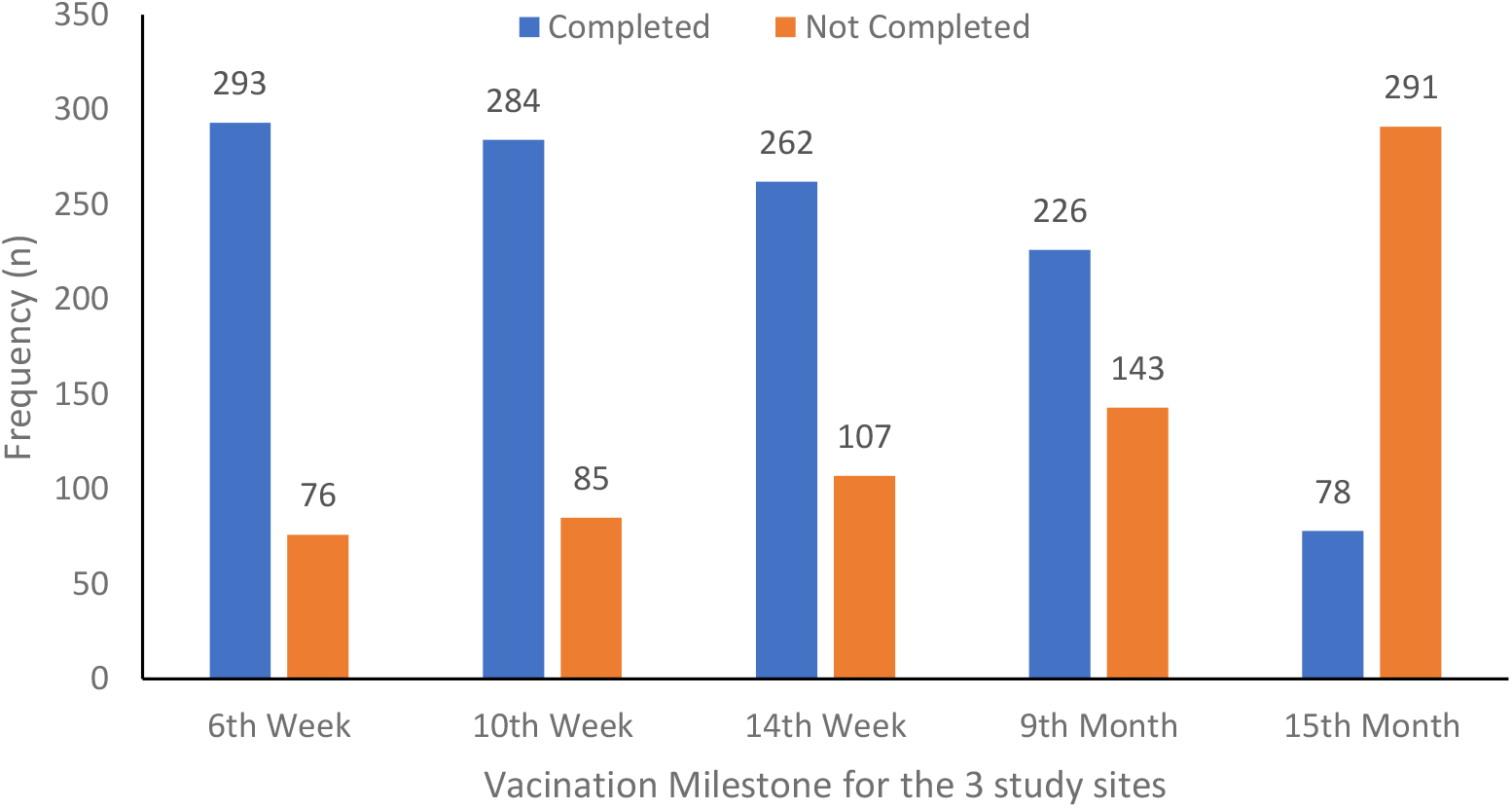
Vaccination completion across immunization milestones in the three study sites

Vaccination coverage across the immunization timeline showed variability at each scheduled visit. At the 6th week, complete vaccination was recorded in 79.7% (98/123) of children in LGA A, 68.3% (84/123) in LGA B, and 90.2% (111/123) in LGA C, while incomplete vaccination accounted for 20.3%, 31.7%, and 9.8%, respectively (Table 3 & Figure 4). The difference across LGAs was statistically significant (χ^2^ = 18.12, p = 0.0001). At the 10th week, complete vaccination coverage was 78.0% (96/123) in LGA A, 70.7% (87/123) in LGA B, and 82.1% (101/123) in LGA C, with incomplete vaccination rates of 22.0%, 29.3%, and 17.9%, respectively. The observed differences were not statistically significant (χ^2^ = 4.61, p = 0.099). At the 14th week, 71.5% (88/123) of children in LGA A, 61.0% (75/123) in LGA B, and 80.5% (99/123) in LGA C had complete vaccination, while incomplete vaccination was reported in 28.5%, 39.0%, and 19.5%, respectively. These differences were statistically significant (χ^2^ = 11.39, p = 0.003). At the 9th month, complete vaccination coverage declined to 59.3% (73/123) in LGA A and 48.0% (59/123) in LGA B but remained higher in LGA C at 76.4% (94/123). Incomplete vaccination was 40.7%, 52.0%, and 23.6%, respectively. The variation across LGAs was statistically significant (χ^2^ = 21.26, p = 0.00001). At the 15th month, complete vaccination was lowest across all LGAs, with 22.0% (27/123) in LGA A, 13.0% (16/123) in LGA B, and 28.5% (35/123) in LGA C. Correspondingly, incomplete vaccination rates were 78.0%, 87.0%, and 71.5%, respectively. The differences remained statistically significant (χ^2^ = 8.87, p = 0.012).

**Table 3.** Vaccination coverage across immunization visits.

| Timeline | Category | LGA A (n = 123), n (%) | LGA B (n = 123), n (%) | LGA C (n = 123), n (%) | $\chi^2$ (p-value) |
| --- | --- | --- | --- | --- | --- |
| 6th week | Complete vaccination | 98 (79.7) | 84 (68.3) | 111 (90.2) | 18.12 (0.0001)* |
|  | Incomplete vaccination | 25 (20.3) | 39 (31.7) | 12 (9.8) |  |
| 10th week | Complete vaccination | 96 (78.0) | 87 (70.7) | 101 (82.1) | 4.61 (0.099) |
|  | Incomplete vaccination | 27 (22.0) | 36 (29.3) | 22 (17.9) |  |
| 14th week | Complete vaccination | 88 (71.5) | 75 (61.0) | 99 (80.5) | 11.39 (0.003)* |
|  | Incomplete vaccination | 35 (28.5) | 48 (39.0) | 24 (19.5) |  |
| 9th month | Complete vaccination | 73 (59.3) | 59 (48.0) | 94 (76.4) | 21.26 (0.00001)* |
|  | Incomplete vaccination | 50 (40.7) | 64 (52.0) | 29 (23.6) |  |
| 15th month | Complete vaccination | 27 (22.0) | 16 (13.0) | 35 (28.5) | 8.87 (0.012)* |
|  | Incomplete vaccination | 96 (78.0) | 107 (87.0) | 88 (71.5) |  |

\*Data are presented as n (%); percentages are calculated within each Local Government Area. *Statistically significant at $p < 0.05$ .*

**Figure 4:**
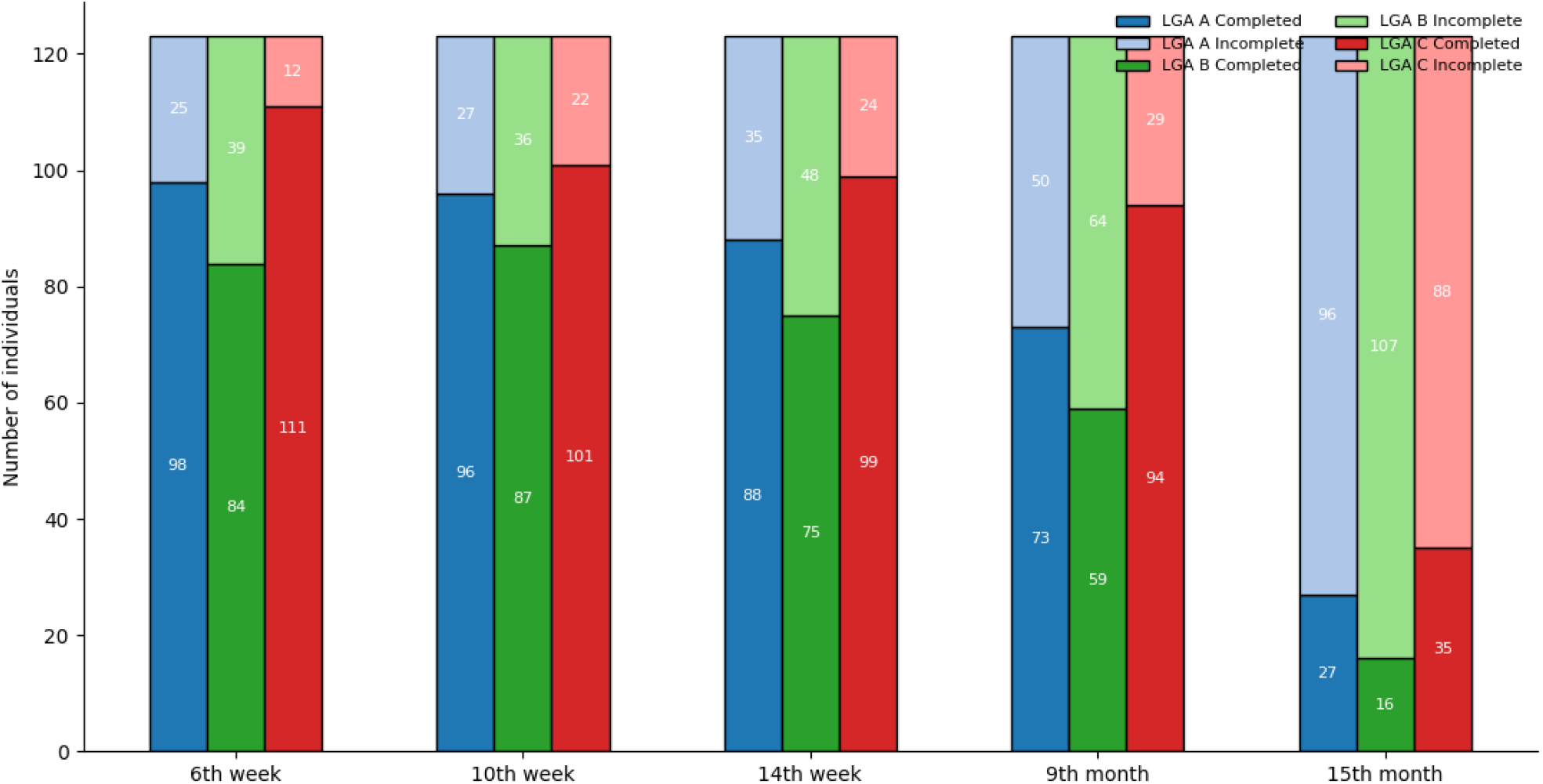
Vaccination coverage across all immunization visits

Figure 5 illustrates the primary reasons for dropout from vaccination across three LGAs (A, B, and C). Among the respondents, forgetting the appointment date was the most frequently reported reason, particularly in LGA B (48 individuals), followed by LGA C (30) and LGA A (34). Other notable reasons included travel (LGA A: 26; LGA C: 22; LGA B: 21) and being busy (LGA A: 20; LGA B: 20; LGA C: 16). Relocation accounted for a smaller proportion of dropouts, ranging from 14 to 15 individuals across the LGAs, while unavailability of health workers was the least common reason, reported by 1-9 individuals. Figure 5 emphasizes that logistical and scheduling challenges are the primary contributors to vaccination discontinuation, highlighting potential areas for targeted intervention to improve adherence.

**Figure 5:**
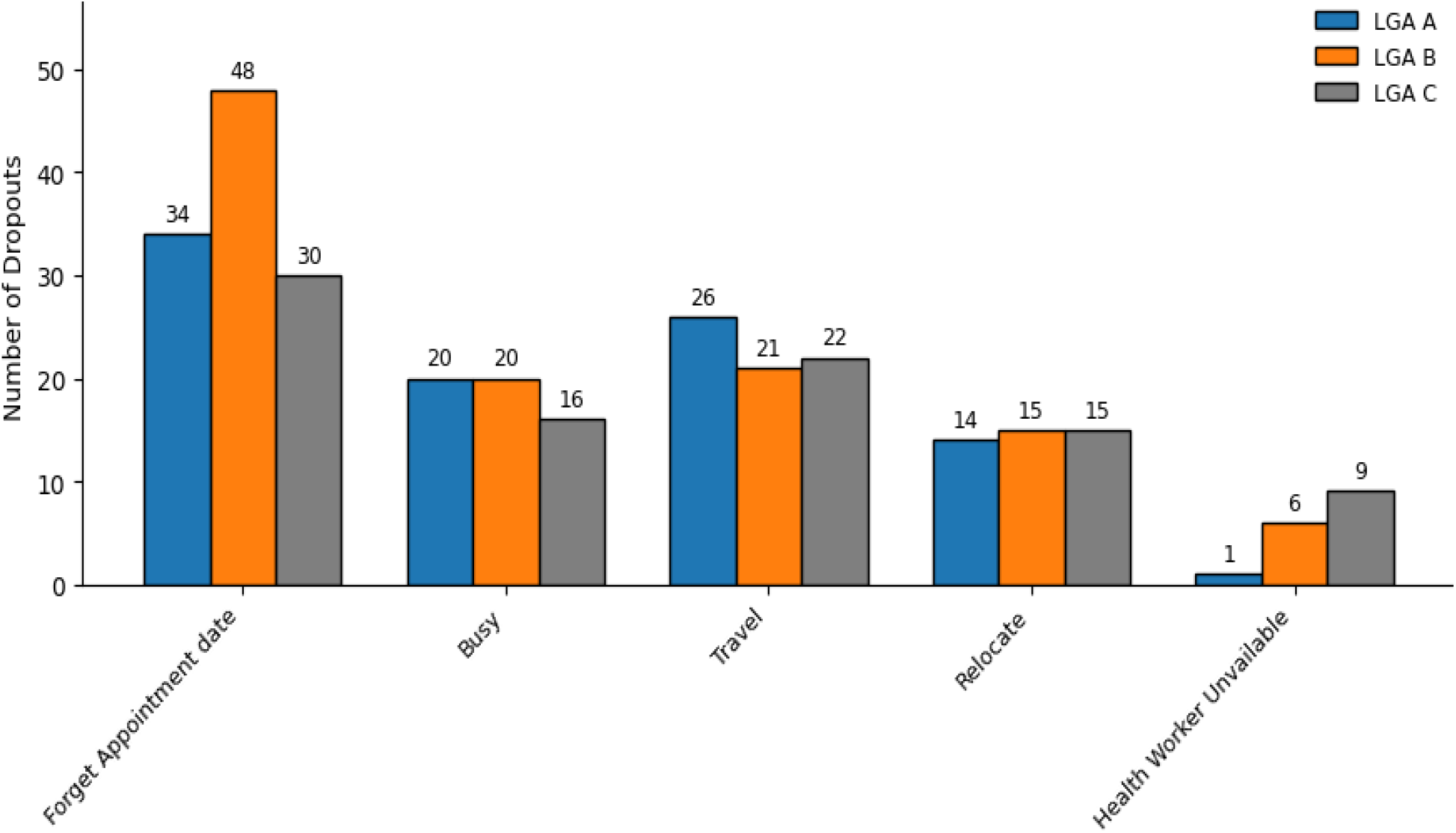
Reasons for vaccination dropout by Local Government Area (LGA)

## 4. Discussion

### 4.1 Sociodemographic Characteristics and Vaccination Uptake

The sociodemographic profile of mother-child pairs in Bayelsa State revealed critical determinants influencing vaccination completion, consistent with prior regional and international studies. Maternal age, education, and occupation demonstrated marked associations with adherence to immunization schedules, mirroring patterns observed in Senegal, Ethiopia, and Togo, where maternal literacy and age were predictive of full immunization (Abdur *et al*., 2019; Lakew *et al*., 2015; Landoh *et al*., 2016; Tsegaw *et al*., 2023; Promise *et al*., 2024; Adelekun *et al*., 2025; Olulaja & Jaber, 2026; Raimi *et al*., 2022). Specifically, secondary education predominated across all LGAs, suggesting that literacy positively influences health-seeking behaviors, as documented in Nigeria and other low-resource settings (Brown & Oluwatosin, 2017; Oyo-Ita *et al*., 2021; Oladepo *et al*., 2021; Eze *et al*., 2018; World Health Organization Africa, 2019a; Chen *et al*., 2016; Domek *et al*., 2019; Promise *et al*., 2026). Occupation further shaped access to vaccination, with business/trading mothers demonstrating higher adherence than farming or informal sector counterparts, echoing findings from prior Bayelsa-based analyses (Adelekun *et al*., 2025; Promise *et al*., 2026; Raimi *et al*., 2022; Olulaja & Jaber, 2026; Kakwi *et al*., 2024a; Oweibia *et al*., 2024; Abdur *et al*., 2019; Lakew *et al*., 2015). Moreover, marital status and household support structures likely facilitated engagement with immunization programs, consistent with evidence from Cross River State and rural Ethiopia (Oyo-Ita *et al*., 2021; Promise *et al*., 2026; Domek *et al*., 2019; Oladepo *et al*., 2021; Eze *et al*., 2018; Chen *et al*., 2016; Abdur *et al*., 2019; Landoh *et al*., 2016). These findings collectively underscore the importance of targeting socioeconomically and educationally disadvantaged populations to enhance vaccine coverage (Raimi *et al*., 2022; Adelekun *et al*., 2025; Promise *et al*., 2026; World Health Organization, 2021; Abdur *et al*., 2019; Olulaja & Jaber, 2026; Brown & Oluwatosin, 2017; Chen *et al*., 2016). In addition, birth order and gender dynamics influenced vaccination completion, aligning with previous observations in sub-Saharan Africa, where first-born children often receive prioritized care due to maternal vigilance (Lakew *et al*., 2015; Landoh *et al*., 2016; Abdur *et al*., 2019; Tsegaw *et al*., 2023; Promise *et al*., 2026; Olulaja & Jaber, 2026; Adelekun *et al*., 2025; Oyo-Ita *et al*., 2021). Conversely, later-born children were more likely to experience delayed or incomplete vaccination, a phenomenon documented in Ethiopia and Togo (Lakew *et al*., 2015; Landoh *et al*., 2016; Tsegaw *et al*., 2023; Promise *et al*., 2026; Abdur *et al*., 2019; Olulaja & Jaber, 2026; World Health Organization Africa, 2019a; Domek *et al*., 2019). This pattern may reflect competing caregiving demands in larger households, reinforcing the need for targeted outreach for multi-child families (Promise *et al*., 2026; Abdur *et al*., 2019; Adelekun *et al*., 2025; Chen *et al*., 2016; Brown & Oluwatosin, 2017; Oladepo *et al*., 2021; Oyo-Ita *et al*., 2021; World Health Organization, 2021). Moreover, the relatively balanced gender distribution in LGA C contrasts with the skewed male-to-female ratios in LGAs A and B, suggesting that contextual sociocultural norms may mediate health-seeking behavior (Raimi *et al*., 2022; Promise *et al*., 2026; Adelekun *et al*., 2025; Abdur *et al*., 2019; Tsegaw *et al*., 2023; Domek *et al*., 2019; Chen *et al*., 2016; Olulaja & Jaber, 2026). Collectively, these sociodemographic factors underscore that interventions must be sensitive to family structure, maternal age, and occupation to optimize vaccine adherence (Brown & Oluwatosin, 2017; Lakew *et al*., 2015; Oyo-Ita *et al*., 2021; Promise *et al*., 2026; World Health Organization, 2021; Abdur *et al*., 2019; Chen *et al*., 2016; Domek *et al*., 2019).

### 4.2 Vaccination Completion and Dropout Patterns

Completion rates were highest in LGA C and lowest in LGA B, illustrating geographic disparities in immunization coverage within Bayelsa State (Adelekun *et al*., 2025; Promise *et al*., 2026; Raimi *et al*., 2022; Kakwi *et al*., 2024a, b). Phone-based reminders consistently outperformed routine follow-up and home visits, confirming the effectiveness of digital interventions in reducing dropout, particularly in hard-to-reach or resource-constrained areas (Chen *et al*., 2016; Domek *et al*., 2019; Mekonnen *et al*., 2021; Promise *et al*., 2026). Attrition along the immunization continuum increased progressively from the 6th week to the 15th month, consistent with global observations of waning adherence over time (Lakew *et al*., 2015; Landoh *et al*., 2016; Tsegaw *et al*., 2023; Promise *et al*., 2026). Early milestones showed higher completion, whereas later stages exhibited dropout exceeding completion, highlighting critical windows for targeted interventions (Adelekun *et al*., 2025; Brown & Oluwatosin, 2017; Mekonnen *et al*., 2021). LGA-specific disparities suggest that local health infrastructure and community engagement influence adherence and that interventions should consider contextual variation (Teddy *et al*., 2025; Raimi *et al*., 2022).

### 4.3 Determinants of Dropout

Analysis of dropout reasons revealed caregiver-level barriers such as forgetfulness, travel, and competing responsibilities, while system-level barriers such as health worker availability contributed minimally. This distinction aligns with prior evidence emphasizing behavioral and logistical constraints as primary determinants of non-completion in sub-Saharan Africa (Lakew *et al*., 2015; Abdur *et al*., 2019; Promise *et al*., 2026; Domek *et al*., 2019; Mekonnen *et al*., 2021). Interventions such as mobile phone reminders, household follow-ups, and community mobilization effectively mitigated dropout, demonstrating that combining behavioral nudges with system support improves completion (Chen *et al*., 2016; Eze *et al*., 2018; Oladepo *et al*., 2021; Promise *et al*., 2026). Attrition analysis indicates that these interventions are particularly critical for later milestones when dropout is highest (Lakew *et al*., 2015; Tsegaw *et al*., 2023; Promise *et al*., 2026).

## 5. Study Limitations

This study has several limitations. First, while the study followed a longitudinal design for immunization milestones, the sample was limited to three LGAs, which may affect generalizability to other regions with different sociocultural or healthcare contexts. Second, reliance on caregiver interviews and immunization registers introduces potential recall, reporting, and documentation biases, particularly in households with multiple children or incomplete records. Third, unmeasured factors such as household mobility, caregiver health literacy, and informal healthcare access could have influenced adherence outcomes. Fourth, variations in outreach intensity, timing, and operational capacity across communities may have affected observed completion rates, potentially underestimating or overestimating true vaccination coverage. Finally, although associations between maternal and child characteristics and vaccination adherence were examined, the observational nature of the study limits causal inference, capturing correlational patterns rather than definitive cause-effect relationships. Despite these constraints, the study provides a detailed overview of immunization adherence dynamics and identifies actionable intervention points across the immunization schedule.

## 6. Summary of the Findings

This study systematically assessed vaccination coverage and adherence among 369 mother-child pairs across three LGAs in Bayelsa State. Sociodemographic factors, including maternal age, education, occupation, and marital status, were significant determinants of immunization completion, with higher literacy and engagement correlating with improved adherence. Completion rates varied geographically, with LGA C achieving the highest coverage and LGA B the lowest, reflecting disparities in access, health infrastructure, and follow-up strategies. Phone-based reminders consistently outperformed routine follow-up and home visits, demonstrating the effectiveness of proactive, scalable communication strategies in reducing dropout. Temporal analysis across immunization milestones showed high early-stage adherence, followed by progressive attrition, particularly by the 15th month. The primary reasons for non-completion were caregiver-level barriers such as forgetfulness, travel, and competing responsibilities, highlighting the interaction of behavioral and logistical constraints with operational factors. Overall, these findings emphasize that vaccination outcomes are shaped by a combination of sociodemographic, behavioral, and system-level factors, and they identify priority areas for interventions, such as targeted caregiver support, milestone-specific outreach, and digital engagement, to enhance adherence and equity in routine immunization programs.

## 7. Implications for Policy and Interventions

The findings of this study have important implications for immunization policy and public health interventions in Bayelsa State and similar contexts.

i. **Targeted support for vulnerable populations:** Interventions should address sociodemographic disparities by enhancing maternal health literacy and providing support for caregivers in rural and low-resource communities, particularly multi-child households.
ii. **Milestone-specific strategies:** Attrition along the immunization schedule highlights the need for focused interventions during later stages, when dropout is most pronounced, to sustain adherence and prevent incomplete vaccination.
iii. **Integration of digital and community engagement tools:** Phone-based reminders and combined home visits effectively reduce behavioral and structural barriers. Incorporating these tools into routine immunization programs can enable scalable, cost-effective engagement.
iv. **Context-sensitive planning:** Policies should consider local health infrastructure, resource allocation, and sociocultural norms to ensure equitable access, consistent service delivery, and tailored outreach responsive to logistical challenges such as forgetfulness, mobility, and competing household responsibilities.

Implementing these strategies can enhance overall immunization coverage, reduce preventable childhood morbidity, and strengthen the resilience and effectiveness of primary healthcare systems.

## 8. Conclusion

Vaccination adherence among children in Bayelsa State is influenced by a combination of sociodemographic, behavioral, and operational factors. Maternal characteristics, including education, age, and occupation, along with proactive strategies such as phone-based reminders and community engagement, significantly enhance completion rates. Geographic disparities, caregiver-level barriers, and logistical constraints contribute to progressive dropout, particularly at later immunization milestones, highlighting critical windows for targeted intervention. These findings underscore the need for sustained, context-specific strategies that integrate digital reminders, milestone-focused outreach, and local health system strengthening to reduce dropout and promote equitable access. By addressing both individual and structural determinants, policymakers and health practitioners can improve routine immunization coverage, enhance child health outcomes, and support progress toward national and global vaccination targets.

## 9. Health Significance

The findings of this study have substantial implications for child health and public health outcomes in Bayelsa State. By identifying key determinants of vaccination adherence, including maternal characteristics and operational factors, the study underscores the critical role of timely and complete immunization in preventing vaccine-preventable diseases. Improved adherence, particularly through phone-based reminders and targeted community engagement, can reduce the incidence of childhood morbidity and mortality, enhance herd immunity, and mitigate outbreaks of preventable infections. Addressing dropout points along the immunization timeline ensures sustained protection during vulnerable periods of early childhood. Moreover, understanding barriers such as forgetfulness, travel, and competing household responsibilities enables the design of context-specific interventions, ultimately strengthening primary healthcare delivery and supporting long-term health equity. These insights provide a foundation for policies and programs that can significantly improve child survival and contribute to achieving broader public health targets. Thus, graphically it is represented (Figure 6 below) as:

**Figure 6:**
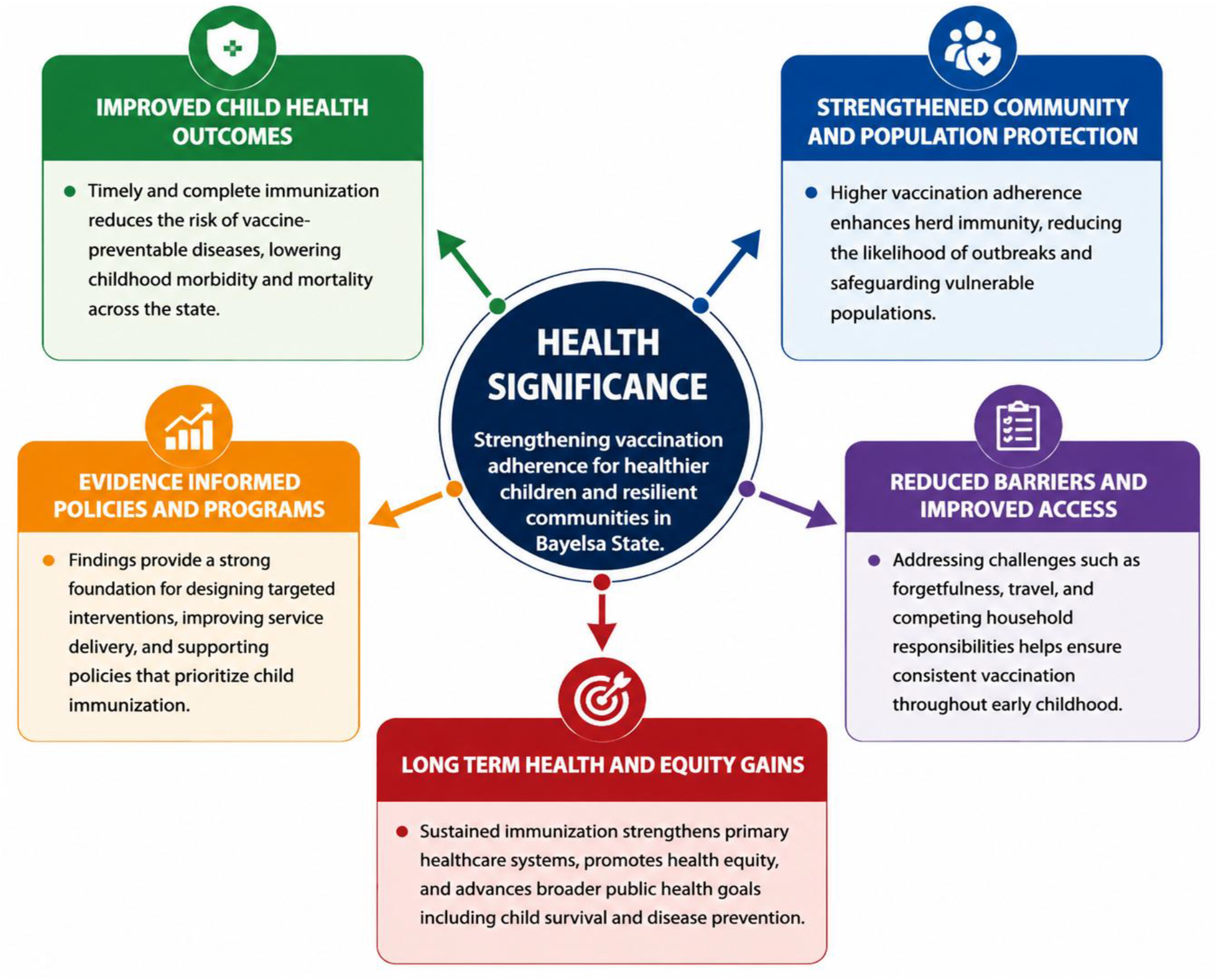
Health Significance of Vaccination Adherence in Bayelsa State, Nigeria: Key Determinants, Outcomes, and Long-Term Public Health Impacts.

## 10. Recommendations

Based on the study findings, the following recommendations are proposed to enhance vaccination coverage and adherence in Bayelsa State, organized into short-term, mid-term, and long-term strategies:

### 10.1 Short-term Recommendations

i. Implement phone-based reminders for upcoming vaccination appointments to reduce missed visits.
ii. Strengthen community outreach during early immunization milestones (6th and 10th weeks) to sustain high initial adherence.
iii. Conduct targeted awareness campaigns emphasizing the importance of completing the full immunization schedule, focusing on caregivers with lower literacy levels.
iv. Mobilize local health workers to provide flexible home visits for families with mobility or scheduling challenges.

### 10.2 Mid-term Recommendations

i. Develop milestone-specific interventions addressing dropout at later stages (14th week, 9th month, and 15th month).
ii. Integrate digital tracking systems in health facilities to monitor completion and identify children at risk of dropout in real time.
iii. Enhance collaboration with community leaders and organizations to support culturally sensitive education and engagement programs.
iv. Train health workers on effective communication and proactive follow-up strategies to reinforce caregiver engagement.

### 10.3 Long-term Recommendations

i. Strengthen health system infrastructure and equitable resource allocation across LGAs to address geographic disparities.
ii. Incorporate vaccination adherence metrics into maternal and child health policies for sustained monitoring and accountability.
iii. Promote community-wide health literacy initiatives, including adult education, to increase understanding of vaccination benefits and reduce behavioral barriers.
iv. Establish ongoing evaluation and research programs to track immunization trends, assess interventions, and inform policy adjustments.

Collectively, these recommendations provide a structured roadmap to improve immunization adherence, reduce preventable childhood morbidity and mortality, and strengthen an equitable and resilient primary healthcare system.

## Data Availability

All data will be made available to interested individuals upon request from the corresponding author

## Transparency

The corresponding author (Morufu Olalekan Raimi) affirms that this manuscript is an honest, accurate, and transparent account of the study being reported; that no important aspects of the study have been omitted; and that any discrepancies from the study as planned (and, if relevant, registered) have been explained.

